# White matter disruption as a cause or consequence of schizophrenia: A Mendelian randomization study

**DOI:** 10.1101/2021.08.23.21262451

**Authors:** Oskar Hougaard Jefsen, Maria Speed, Karl John Friston, Søren Dinesen Østergaard, Doug Speed

**Author notes:** **Corresponding author** Oskar Hougaard Jefsen, Medical Doctor, Psychosis Research Unit, Aarhus University Hospital, Denmark, Palle Juul-Jensens Blvd. 175, Aarhus N.

## Abstract

Schizophrenia is hypothesized to be caused by impaired functional integration in the brain, and this could hypothetically be caused by white matter disruptions, synaptic dysfunction, or both. Neuroimaging studies consistently show reduced fractional anisotropy, a measure of white matter integrity, in patients with schizophrenia. Using Mendelian randomization, we show that these white matter changes are likely to be the consequence of a primary synaptopathy in schizophrenia.

## Main text

Schizophrenia (SZ) is a severe mental illness affecting ∼ 1% of the population. The etiology of SZ is unknown, but likely involves a complex interplay between genetic and environmental risk factors shaping brain development.^1^ Despite its elusive etiology, an overarching idea of schizophrenia as a disintegration of the mind has persisted from the very inception of schizophrenia as a diagnostic construct till present day. The disintegration-idea comes in two versions (Figure 1). One version implies that functional integration is impaired due to anatomical disruptions in white matter fiber tracts, causing *dis*connection between regions (“dis” = “apart”) and can be traced back to Wernicke’s “sejunction hypothesis”.^2^ The other version implies that functional integration is impaired due to changes at the level of synaptic efficacy and plasticity, causing *dys*connection between brain regions (“dys” = “ill”), an idea more in line with Bleuler’s conception of schizophrenia.^3^ Recently, modern neuroimaging and connectivity analyses have provided novel empirical evidence for impaired functional integration in schizophrenia, however, whether these impairments are caused by anatomical disruptions or by synaptic dysfunction remains unresolved.

**Figure 1.**
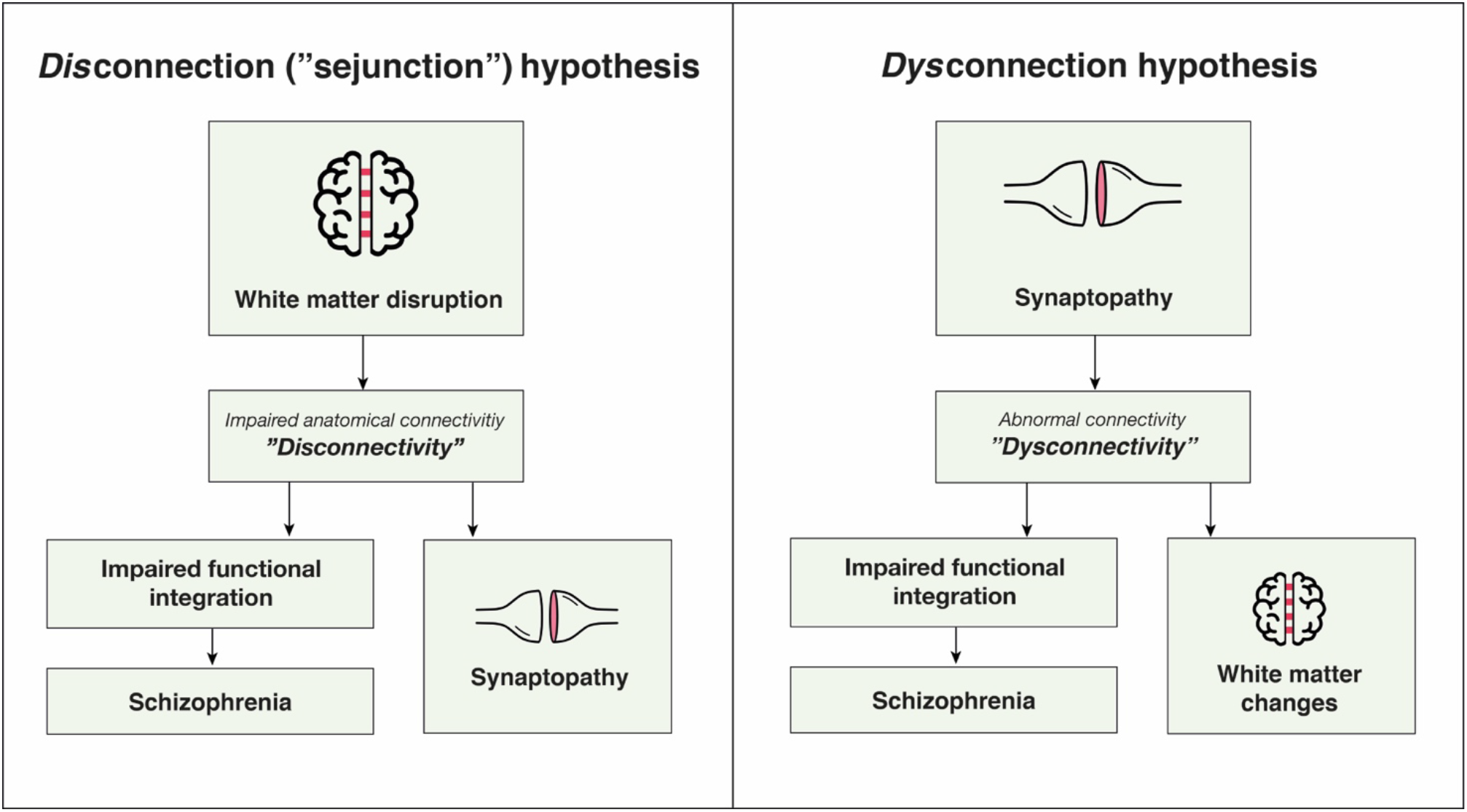
Graphical representation of two hypotheses to explain the role of white matter disruption in schizophrenia.

In favor of an anatomical *dis*connection hypothesis, diffusion tensor imaging (DTI) studies on patients with SZ have found reduced fractional anisotropy (FA) in cerebral white matter tracts, particularly in the corpus callosum.^4-6^ In white matter, FA is primarily affected by fiber orientation, myelination, and axonal density, and can cautiously be interpreted as a measure of white matter integrity.^7^ In favor of a synaptic *dys*connection hypothesis, several lines of evidence point towards synaptic dysfunction as the primary cause of schizophrenia,^8,9^ e.g. studies in computational neuroscience,^10,11^ pharmacology,^12^, and genetics.^13^ Specifically, genome-wide association studies (GWAS) of schizophrenia have revealed strong enrichment for genes implicated in synaptic organization, differentiation, and transmission,^13^ pointing away from a primary anatomical cause of schizophrenia. In this view, the anatomical (white matter) changes are seen as secondary consequences to a primary synaptic dysfunction (Figure 1).^9^ Intriguingly, a prior study found a negative association between a polygenic risk score (PRS) for schizophrenia and white matter FA,^14^ but this correlational analysis does not allow for causal inference.

In summary, the *dys*connection hypothesis attributes the primary pathophysiology to a synaptopathy (particularly involving the modulation of synaptic gain and efficacy) that leads to secondary anatomical *dis*connection (via experience-dependent plasticity, synaptic regression, and accompanying activity-dependent myelination). Conversely, the “sejunction hypothesis” considers white matter disruption (leukodystrophy) as the primary (e.g., neurodevelopmental, or epigenetic) etiology, leading to a secondary synaptopathy that underwrites schizophrenia.

In the present study, we aimed to infer the causal relationship between schizophrenia and FA using Mendelian randomization, a method from genetic epidemiology that uses genetic variants (single nucleotide polymorphisms, SNPs) as instrumental variables for heritable traits of interest.^15^ In brief, Mendelian randomization licenses causal inference by using instrumental variables (based upon SNPs) to relax the assumption of independence between exposure (e.g., schizophrenia) and unmodelled (e.g., random) effects on outcomes (e.g., FA reductions). Our analyses used GWAS summary statistics from the Oxford Brain Imaging Genetics (BIG40)^16^ web server and the Schizophrenia Working Group of the Psychiatric Genomics Consortium.^17^ The BIG40 web server contains summary statistics from ∼ 4,000 image-derived phenotypes (IDPs) measured in almost ∼ 40,000 individuals from the UK Biobank. We focused our investigation on FA in the corpus callosum for two reasons: First, because reduced FA in the corpus callosum is a consistent finding in schizophrenia across studies^4-6^ and second, because the corpus callosum has a strict left-right orientation, eliminating fiber orientation as a confound of FA.^7^

Using Mendelian randomization^18-20^ (see Online methods^19,20^), we found nominally significant evidence for a causal effect of (genetic liability to) schizophrenia on FA reductions in the genu (logOR=-0.028, SD=0.13, *p*=0.037) and splenium (logOR=-0.025, SD=0.13, *p*=0.047) of the corpus callosum, but not the body of the corpus callosum (logOR=-0.020, SD=0.013, *p*=0.106). As a sensitivity analysis, we performed Egger regression, and found no evidence of pleiotropy. We chose not to adjust for multiple comparisons due to the non-independence between comparisons (the genetic correlations between pairs of FA measures were between 0.65 and 0.88). However, if one does apply Bonferroni-correction for the three comparisons, none of the tests are statistically significant (α = .05), underscoring the weakness of the presented evidence.

Conversely, we found no evidence of a causal effect of reduced FA on schizophrenia in any part of the corpus callosum. This negative finding must, of course, be interpreted in light of the relatively weaker genetic instrument for FA in the corpus callosum (explaining 2.4-3% of the variance), compared to the slightly stronger genetic instrument for schizophrenia (explaining 6.4% of the variance). Importantly in this regard, a causal effect of genetic liability to schizophrenia on white matter disruption, does not preclude a contributory causal role of white matter disruption on schizophrenia.

In summary, our findings provide preliminary evidence that white matter disruptions may be a consequence of a primary synaptopathy in schizophrenia. We can, however, not draw firm conclusions on this question due to the small effect sizes, and consider them merely to represent putative evidence for the primacy of synaptic, over anatomical, disruptions in schizophrenia. We propose that this step be followed by replication studies and other lines of investigation, to further the understanding of the etiology of schizophrenia.

## Data Availability

Not applicable.

## Acknowledgements

O.H.J. is supported by a grant from the Health Research Foundation of Central Denmark Region. D.S. is supported by Aarhus University Research Foundation (AUFF), by the Independent Research Fund Denmark under Project no. 7025-00094B, and by a Lundbeck Foundation Experiment Grant.

## Author contributions

O.H.J. conceived of the study, interpreted the results, and drafted the first version of the manuscript. M.S. and D.S. obtained the summary statistics, analyzed the data, interpreted the results, and revised the manuscript for important intellectual content. K.J.F. revised the manuscript for important intellectual content. S.D.Ø. contributed to the study design, interpreted the results, and revised the manuscript for important intellectual content. All authors approved the final version of the manuscript prior to submission.

---

**Table 1.**
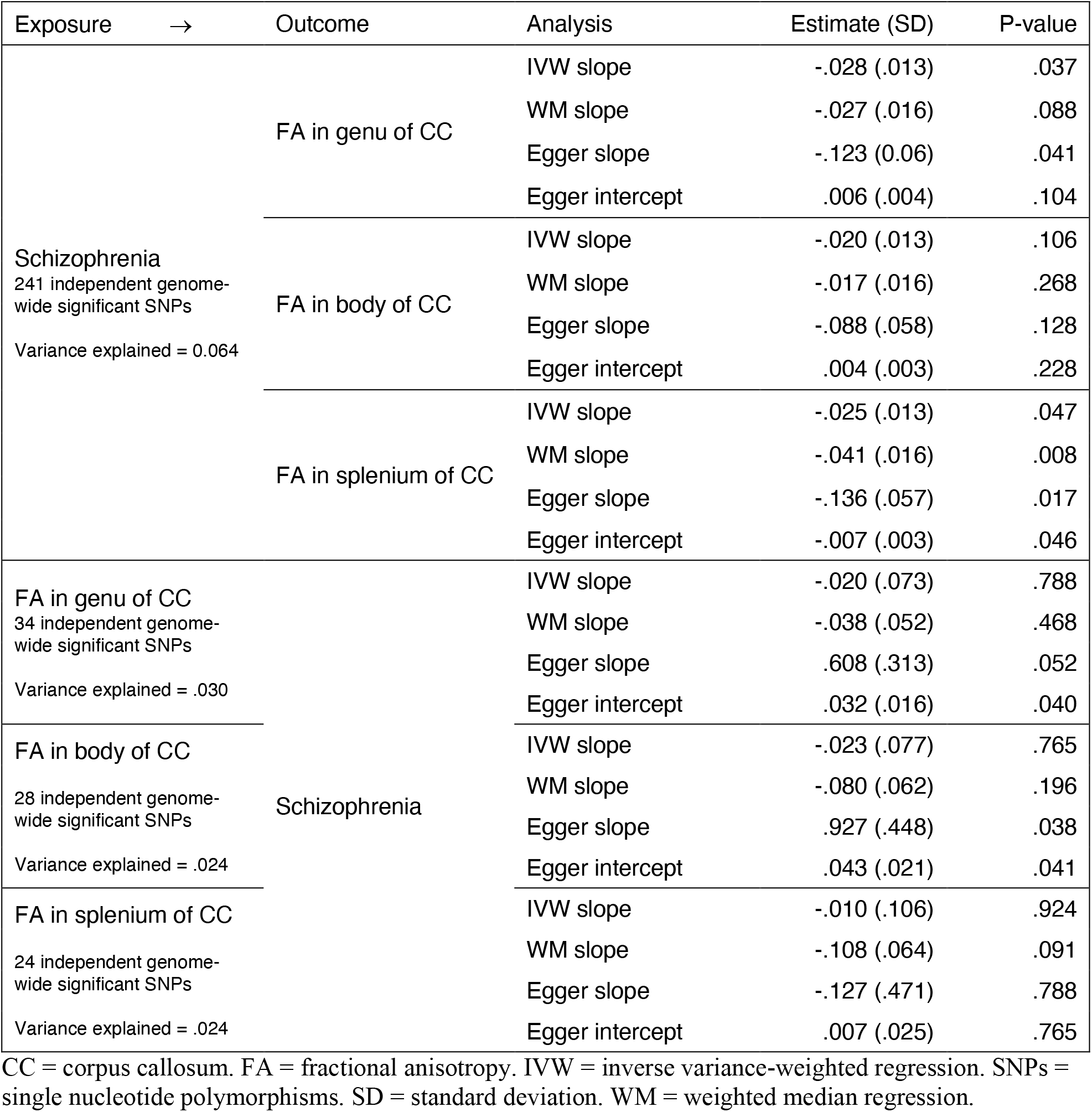

## References

1 Kahn, R. S. et al.. Schizophrenia. Nature Reviews Disease Primers 1, 15067, doi:10.1038/nrdp.2015.67 (2015).

2 Wernicke, C. Grundriss der Psychiatrie in klinischen Vorlesungen. (Thieme, 1906).

3 Bleuler, E. Dementia praecox or the group of schizophrenias New York, NY International Universities Press (1911 [in German]).

4 Ellison-Wright, I. & Bullmore, E. Meta-analysis of diffusion tensor imaging studies in schizophrenia. Schizophrenia research 108, 3–10 (2009).

5 Kelly, S. et al.. Widespread white matter microstructural differences in schizophrenia across 4322 individuals: results from the ENIGMA Schizophrenia DTI Working Group. Molecular psychiatry 23, 1261–1269 (2018).

6 Koshiyama, D. et al.. White matter microstructural alterations across four major psychiatric disorders: mega-analysis study in 2937 individuals. Molecular psychiatry 25, 883–895 (2020).

7 Friedrich, P. et al.. The relationship between axon density, myelination, and fractional anisotropy in the human Corpus callosum. Cerebral Cortex 30, 2042–2056 (2020).

8 Friston, K. J. & Frith, C. D. Schizophrenia: a disconnection syndrome. Clin Neurosci 3, 89–97 (1995).

9 Friston, K., Brown, H. R., Siemerkus, J. & Stephan, K. E. The dysconnection hypothesis (2016). Schizophrenia research 176, 83–94 (2016).

10 Bastos-Leite, A. J. et al.. Dysconnectivity within the default mode in first-episode schizophrenia: a stochastic dynamic causal modeling study with functional magnetic resonance imaging. Schizophrenia bulletin 41, 144–153 (2015).

11 Adams, R. A. et al.. Computational modelling of EEG and fMRI paradigms indicates a consistent loss of pyramidal cell synaptic gain in schizophrenia. Biological Psychiatry (article in press), doi:10.1016/j.biopsych.2021.07.024 (2021).

12 Corlett, P. R., Honey, G. D., Krystal, J. H. & Fletcher, P. C. Glutamatergic model psychoses: prediction error, learning, and inference. Neuropsychopharmacology 36, 294–315 (2011).

13 Ripke, S., Walters, J. T., O’Donovan, M. C. & Consortium, S. W. . o. t. P. G. GMapping genomic loci prioritises genes and implicates synaptic biology in schizophrenia. MedRxiv (2020).

14 Reus, L. M. et al.. Association of polygenic risk for major psychiatric illness with subcortical volumes and white matter integrity in UK Biobank. Scientific reports 7, 1–8 (2017).

15 Davey Smith, G. & Ebrahim, S. ‘Mendelian randomization’: can genetic epidemiology contribute to understanding environmental determinants of disease? International journal of epidemiology 32, 1–22 (2003).

16 Smith, S. M. et al.. An expanded set of genome-wide association studies of brain imaging phenotypes in UK Biobank. Nature neuroscience 24, 737–745 (2021).

17 Ripke, S. et al.. Biological insights from 108 schizophrenia-associated genetic loci. Nature 511, 421 (2014).

18 Yavorska, O. O. & Burgess, S. MendelianRandomization: an R package for performing Mendelian randomization analyses using summarized data. International journal of epidemiology 46, 1734–1739 (2017).

19 Burgess, S., Bowden, J., Fall, T., Ingelsson, E. & Thompson, S. G. Sensitivity analyses for robust causal inference from Mendelian randomization analyses with multiple genetic variants. Epidemiology (Cambridge, Mass.) 28, 30 (2017).

20 Bowden, J. et al.. Assessing the suitability of summary data for two-sample Mendelian randomization analyses using MR-Egger regression: the role of the I2 statistic. International journal of epidemiology 45, 1961–1974 (2016).

