## Supplementary methods for "White matter disruption as a cause or consequence of schizophrenia: A Mendelian randomization study"

**Affiliations**
[1] Psychosis Research Unit, Aarhus University Hospital – Psychiatry, Denmark.
[2] Department of Clinical Medicine, Aarhus University, Aarhus, Denmark.
[3] Department of Affective Disorders, Aarhus University Hospital – Psychiatry, Denmark.
[4] The Wellcome Centre for Human Neuroimaging, University College London, UK.
[5] Center for Quantitative Genetics and Genomics, Aarhus University, Aarhus, Denmark

**Keywords**
Schizophrenia, white matter, fractional anisotropy, Mendelian Randomization

**Corresponding author**
Oskar Hougaard Jefsen
Medical Doctor
Psychosis Research Unit, Aarhus University Hospital, Denmark
Palle Juul-Jensens Blvd. 175, Aarhus N


**Genome-wide association study summary statistics**

We used summary statistics from two large GWASs; the latest meta-analysis of schizophrenia by the Schizophrenia Working Group of the Psychiatric Genomics Consortium (PGC) [1], and the GWAS on image-derived phenotypes (IDPs) from magnetic resonance imaging (MRI) in UK Biobank (UKBB) participants.[2]

*Schizophrenia*

In total, the PGC meta-analysis used data from 69,369 schizophrenia cases and 236,642 controls. We downloaded summary statistics for the discovery cohort (67,390 cases and 94,015 controls) from https://www.med.unc.edu/pgc/download-results. Approximately 80% of individuals were of European ancestry.

*Fractional anisotropy in the corpus callosum*

The study by Smith et al. performed GWASs using 39,691 UKBB individuals, divided into a discovery cohort of 22,138 individuals (53% females) and a replication cohort of 11,086 individuals (52% females). The age distribution (mean±SD) in the discovery cohort was 63.7±7.3 years for females, and 65.0±7.6 years for males. The UKBB data included six MRI modalities; here we only used data from diffusion MRI (dMRI). Comprehensive information on the UKBB MRI acquisition and processing pipeline is available in the UKBB brain imaging documentation: https://biobank.ctsu.ox.ac.uk/showcase/showcase/docs/brain_mri.pdf. We downloaded summary statistics for three IDPs from https://open.win.ox.ac.uk/ukbiobank/big40. These IDPs correspond to fractional anisotropy (FA) in three parts of the corpus callosum: genu (IDP_dMRI_TBSS_FA_Genu_of_corpus_callosum, Pheno = 1454, UKBB ID = 25058), body (IDP_dMRI_TBSS_FA_Body_of_corpus_callosum, Pheno = 1455, UKBB ID = 25059), and splenium (IDP_dMRI_TBSS_FA_Splenium_of_corpus_callosum, Pheno = 1456, UKBB ID = 25059).

*Quality control*

First we filtered the summary statistics for schizophrenia. We excluded SNPs with minor allele frequency (MAF) < 0.01, info score < 0.9, sample size ≦ 150,000, that had ambiguous alleles (A&T or C&G) or that were not consistent with the SNPs in 1000 Genome Project.[3] Then we filtered the summary statistics for the three IDPs. We excluded SNPs with MAF < 0.01, ambiguous alleles or that were not consistent with the SNPs in 1000 Genome Project[3] (we did not filter based on info scores or per-SNP samples sizes as these were not available). After these filterings, there were 4,071,224 SNPs common to the two sets of summary statistics.

**Mendelian Randomization analyses**

Mendelian Randomization (MR) is a method for testing whether an exposure is causal for an outcome. In total, we performed six MR analyses. The first three used schizophrenia as the exposure, then one of the IDPs as the outcome; the last three used one of the IDPs as the exposure, then schizophrenia as the outcome. For each analysis, we identified independent genetic instrumental variables for the exposure. To do this, we used the summary statistics for the exposure to identify SNPs that are genome-wide significant ($p < 5 \times 10^{-8}$), then used the 1000 Genome Data to clump SNPs so that no pair remained within 3 centiMorgans with correlation-squared $r^2 > 0.05$.

We performed the Mendelian Randomization analyses using the R-package "Mendelian-Randomization".[4] For the main analysis, we used inverse-variance weighted (IVW) regression to estimate the regression slope (a significant non-zero slope indicates that the exposure is causal for the outcome). We performed weighted-median regression[5], which gives unbiased of the slope provided at least half the SNPs are valid instrumental variables. Furthermore, we performed Egger regression, which tests for the presence of pleiotropic effects.[6] A significant non-zero Egger intercept indicates a direct effect of the instrumental variables (SNPs) onto the outcome, and the slope estimates the remaining, indirect, effect.

### References


1 Ripke, S. *et al.* Biological insights from 108 schizophrenia-associated genetic loci. *Nature* **511**, 421 (2014).

2 Smith, S. M. *et al.* An expanded set of genome-wide association studies of brain imaging phenotypes in UK Biobank. *Nature neuroscience* **24**, 737-745 (2021).

3 Consortium, G. P. An integrated map of genetic variation from 1,092 human genomes. *Nature* **491**, 56-65 (2012).

4 Yavorska, O. O. & Burgess, S. MendelianRandomization: an R package for performing Mendelian randomization analyses using summarized data. *International journal of epidemiology* **46**, 1734-1739 (2017).

5 Burgess, S., Bowden, J., Fall, T., Ingelsson, E. & Thompson, S. G. Sensitivity analyses for robust causal inference from Mendelian randomization analyses with multiple genetic variants. *Epidemiology (Cambridge, Mass.)* **28**, 30 (2017).

6 Bowden, J. *et al.* Assessing the suitability of summary data for two-sample Mendelian randomization analyses using MR-Egger regression: the role of the I2 statistic. *International journal of epidemiology* **45**, 1961-1974 (2016).
